# Wastewater SARS-CoV-2 RNA Concentration and Loading Variability from Grab and 24-Hour Composite Samples

**DOI:** 10.1101/2020.07.10.20150607

**Authors:** Kyle Curtis, David Keeling, Kathleen Yetka, Allison Larson, Raul Gonzalez

## Abstract

The ongoing COVID-19 pandemic caused by severe acute respiratory syndrome coronavirus 2 (SARS-CoV-2) requires a significant, coordinated public health response. Assessing case density and spread of infection is critical and relies largely on clinical testing data. However, clinical testing suffers from known limitations, including test availability and a bias towards enumerating only symptomatic individuals. Wastewater-based epidemiology (WBE) has gained widespread support as a potential complement to clinical testing for assessing COVID-19 infections at the community scale. The efficacy of WBE hinges on the ability to accurately characterize SARS-CoV-2 RNA concentrations in wastewater. To date, a variety of sampling schemes have been used without consensus around the appropriateness of grab or composite sampling. Here we address a key WBE knowledge gap by examining the variability of SARS-CoV-2 RNA concentrations in wastewater grab samples collected every 2 hours for 72 hours compared with three corresponding 24-hour flow-weighted composite samples collected over the same period. Results show relatively low variability (respective means for N1, N2, N3 assays = 608, 847.9, 768.4 copies 100 mL^-1^, standard deviations = 501.4, 500.3, 505.8 copies 100 mL^-1^) for grab sample concentrations, and good agreement between most grab samples and their respective composite (mean deviation from composite = 159 copies 100 mL^-1^). When SARS-CoV-2 RNA concentrations are used to calculate viral load (RNA concentration * total influent flow the sample day), the discrepancy between grabs (log_10_ range for all grabs = 11.9) or a grab and its associated 24-hour composite (log_10_ difference = 11.6) are amplified. A similar effect is seen when estimating carrier prevalence in a catchment population with median estimates based on grabs ranging 63-1885 carriers. Findings suggest that grab samples may be sufficient to characterize SARS-CoV-2 RNA concentrations, but additional calculations using these data may be sensitive to grab sample variability and warrant the use of flow-weighted composite sampling. These data inform future WBE work by helping determine the most appropriate sampling scheme and facilitate sharing of datasets between studies via consistent methodology.

## 1. Introduction

The outbreak of the novel severe acute respiratory syndrome coronavirus 2 (SARS-CoV-2) in late 2019 escalated to a global pandemic. To date (11-2-2020) there are over 46.7 million confirmed cases and 1.2 million deaths world-wide attributed to COVID-19, the disease caused by SARS-CoV-2 (Dong et al., 2020). Understanding the extent and density of infection is critical in effectively responding to this pandemic. However, due to limited diagnostic testing (Babiker et al., 2020, Schneider 2020) and inconsistent reporting of results (Leonor, 2020), generating reliable COVID-19 prevalence estimates in a community remains challenging. This is compounded by asymptomatic disease transmission, the rate of which is still unclear (Kronbichler et al., 2020).

Wastewater-based epidemiology (WBE) represents a promising complement to clinical testing as a means of assessing COVID-19 trends and prevalence within a community. WBE has been used to investigate occurrence and trends for a variety of chemical (pharmaceuticals, illicit drugs) and biological (pathogens, antibiotic resistance genes) constituents at the community-scale by measuring biomarkers in wastewater (Choi et al., 2018, Been et al., 2014, Sims and Ksprzyk-Hordern, 2020). Unlike clinical testing data, which is limited by test availability and biased towards the detection of symptomatic individuals, WBE yields a community-scale viral load estimate for a wastewater treatment facility’s catchment population. Considering these benefits, there has been much support for WBE as a complementary strategy to clinical testing in response to the SARS-CoV-2 pandemic (Ahmed et al., 2020, Bivins et al., 2020, Hata et al., 2020).

The use of WBE in a variety of geographically and demographically disparate areas creates the opportunity to coordinate efforts, assimilate data, and assess SARS-CoV-2 trends on a larger scale than any single WBE study could alone. For this broad, integrated approach to succeed many knowledge gaps must first be addressed for appropriate data comparisons. Such areas include sample collection, preservation, concentration, and quantification in a complex and challenging wastewater matrix (Bivins et al., 2020, Daughton, 2020, Kitajima et al., 2020, Mao et al., 2020, Worley-Morse et al., 2019). A fundamental study design knowledge gap considers how to collect a sample that is appropriately representative of SARS-CoV-2 concentrations in wastewater. Given that influent flows at wastewater facilities fluctuate continually it is important to understand if these variations in flow correspond to significant virus concentration variation. Specifically, do grab samples sufficiently characterize wastewater SARS-CoV-2 concentrations, or are flow-weighted composites necessary?

We address this knowledge gap via a comparison of grab and 24-hr flow-weighted composite samples over a 3-day intensive time series. The goal was to characterize SARS-CoV-2 variability in grab samples collected every 2hrs for 72 hours and compare this variability with 3 flow-weighted composites collected over the same time frame. Specific objectives are; 1) to examine the variability of reverse transcription droplet digital PCR (RT-ddPCR) quantified SARS-CoV-2 RNA concentrations, 2) compare daily loading calculations from grab sample concentrations with loading calculations using respective 24-hr flow-weighted composite concentrations, and 3) compare carrier prevalence estimates from grab sample concentrations with carrier prevalence estimates using respective 24-hr flow-weighted composite concentrations.

This work will aid future WBE studies in determining the most appropriate sampling scheme. Increasing the chance of accurately characterizing SARS-CoV-2 concentrations in wastewater allows WBE work to provide the best available data for use in subsequent calculations, such as estimates of community infections or epidemiological models.

## 2. Methods

### 2.1 Wastewater Treatment Facility

Army Base Treatment Plant (ABTP) is in Norfolk, VA, and is operated by Hampton Roads Sanitation District (HRSD). It services an area of approximately 21 square miles, which is dominated by residential development, a port, and a large military base. The treatment plant serves a population of approximately 78,322, however this figure can fluctuate considerably due to the arrival and departure of military vessels and cargo ships. The catchment population can also vary based on redirection of flow throughout the collection system, a practice that is common for wastewater utilities.

For ABTP, pretreatment involves coarse screening via bar screens. Residual suspended solids, fats, oils, and grease are removed during a primary settling step. Secondary treatment consists of a 5-stage Bardenpho system and secondary settling. Secondary clarifier effluent is disinfected with sodium hypochlorite and dechlorinated via sodium bisulfite prior to discharge. ABTP has a design flow of 68 million liters per day with a peak capacity of 136 million liters per day, and average daily flows ranging 38-42 million liters per day. Over the three-day study period, the average daily flow was 47.2 million liters per day.

### 2.2 Study Design

Samples were aseptically collected over a 72-hour period (5/1/2020 10:00 EST– 5/4/2020 10:00 EST) from the ABTP Raw Water Influent (RWI) sample point prior to pretreatment. Thirty-six uniform 1L grab samples were collected every two hours using an ISCO Avalanche portable refrigerated sampler (Teledyne ISCO, Lincoln, NE) which kept the samples at approximately 4°C. For each of 3, 24-hour periods, a flow-weighted composite sample was collected concurrently with the sequentially collected grabs using an ISCO 3710 Portable sampler (Teledyne ISCO). The composite sampler was paced to take a 150mL aliquot every 870,550 liters, with all aliquots collected in a sterile 15L carboy in a sampler base filled with ice that was replenished daily. Final Effluent (FNE) samples were collected aseptically after the 30-minute chlorine contact point between mid-morning and mid-day of each collection. Each set of 24-hour composite samples were transported on ice from the sampling site to the HRSD Central Environmental Laboratory (within 4 hours) where samples were processed upon arrival.

### 2.3 Sample Processing

Electronegative filtration, following the method in Worley-Morse et al. (2019), was used to concentrate SARS-CoV-2 from 50 mL of raw wastewater and 200 mL of treated final effluent. Filters were stored in a - 80°C freezer immediately after concentration until RNA extraction using the NucliSENS easyMag (bioMerieux Inc., Durham, NC, USA) modified protocol described in Worley-Morse et al. Prior to extraction, 1 × 10 copies of Hep G Armored RNA (Asuragen, Austin, TX, USA) was spiked in the lysis buffer for all samples and negative extraction controls to quantify matrix inhibition. One-step RT-ddPCR was used to enumerate SARS-CoV-2 N1, N2, and N3 assays (Lu et al., 2020) and the Hep G inhibition control on a Bio-Rad QX200 (Bio-Rad, Hercules, CA, USA) using the protocol in Gonzalez et al (2020). Inhibition was determined by calculating the Hep G recovery in the samples compared to the NEC. Samples were not re-run when inhibition was an issue (less than 10% recovery). A detailed RT-ddPCR protocol and run summary statistics can be found in the Supplemental Information.

### 2.4 Estimating SARS-CoV-2 Infections in the Sewage Collection System

A promising extension of WBE is calculating prevalence estimates to better gauge the number of infected individuals (both symptomatic and asymptomatic) in a community. This approach has been used in several recent SARS-CoV-2 publications (Ahmed et al., 2020, Medema et al., 2020, Wu et al., 2020). The number of SARS-CoV-2 infected carriers for the ABTP service area were estimated using two values—viral load per person and total viral load to the treatment facility. For the purpose of viral load and carrier prevalence estimates, only the N2 assay was used. N2 was selected based on superior performance in previous work (Gonzalez et al. 2020), assessed by frequency of detection in wastewater.

Equation 1 was used to calculate the viral load per person (the total amount of virus shed by an infected person via feces). The 90^th^ percentile concentration of SARS-CoV-2 in stool reported from Wölfel et al. (2020) was used as variable A in equation 1. A triangular distribution (minimum= 51, likeliest= 128, maximum= 796) for the fecal mass per person per day, variable B, was fitted from Rose et al. (2015). This distribution was sampled during each of 10,000 Monte Carlo simulations conducted using Oracle Crystal Ball (Oracle, Berkshire, UK).

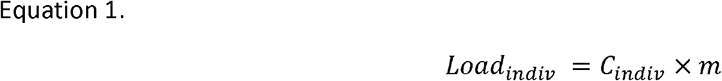

where;

*Load*_*indiv*_ = Viral load per person (copies day^-1^)

*C*_*indiv*_ = concentration of SARS-CoV-2 virus in feces (copies g^-1^)

*m* = typical mass of stool produced per person per day (g day^-1^)

Total viral load to the WWTP during each sampling event was calculated using equation 2. In order to quantify potential carriers in the population the N2 assay concentration for each sample was used as the *C*_*WWTP*_ value in Equation 2.

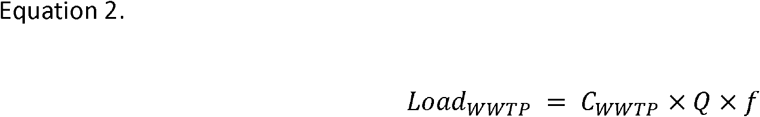

where;

*Load*_*WWTP*_ = Viral load to WWTP (copies day^-1^)

*C*_*WWTP*_ = concentration of SARS-CoV-2 in wastewater samples (copies 100 mL^-1^)

*Q* = Plant flow (million liters per day, million liters day^-1^)

*f* = Conversion factor between 100 mL and million liters day^-1^

Prevalence estimates were calculated using equation 3, which incorporated results from equations 1 and 2 for each sampling event.

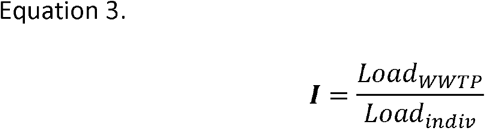

where;

*I* = Estimated proportion of WWTP service area infected

### 2.5 Data Analysis and Visualization

Data analysis and visualization was conducted using R Statistical Computing Software version 3.6.3. (R Core Team, 2019). The dplyr (Wickham et al., 2015) and tidyr (Wickham et al., 2018) packages were primarily used for data manipulation and the ggplot2 package (Wickham 2016) was used for all plotting. The code used to create each figure can be found at https://github.com/mkc9953/WW_EPI_grab_composite_study.

## 3. Results and Discussion

Three large wastewater facilities collect and treat portions of the city of Norfolk’s wastewater. The ABTP currently receives wastewater from approximately 36% of the city’s population. During the study period there was 211, 211, and 239 cumulative clinically confirmed COVID-19 cases in the entire city (for days 1, 2, and 3, respectively). Gonzalez et al. (2020) have been monitoring this facility, amongst others, weekly since March 9^th^, 2020. Detections of SARS-CoV-2 began on April 6, 2020—4 weeks prior to this study.

### 3.1 Influent Flow and Rainfall

Hourly wastewater influent flow during the study period ranged from 27.1 to 61.6 million liters per day, with a mean flow of 46.6 million liters per day and standard deviation of 103 million liters per day. A description of flow characteristics by sample day can be found in Table 1. Two days prior to the first sampling event there was a storm generating approximately 2.5 cm of rainfall. A brief increase in flow was observed, likely due to stormwater infiltrating the sewer collection system. Influent flow at the treatment facility returned to typical dry weather values in approximately 6hrs and remained at levels typical of dry weather throughout the study. No rainfall occurred in the vicinity of the treatment facility during the study period. The treatment facility serves several low-lying areas that are subject to inundation during moderate high tide events, causing saltwater intrusion into sewer collection system. Treatment plant influent conductivity, used as an indicator of seawater, begins to increase significantly following tidal levels greater than 1.07 meters Mean Lower Low Water (MLLW). High tides during the period sampled were 1.04 meters MLLW or less based on the Sewell’s Point Tide Gage operated by NOAA.

**Table 1.**
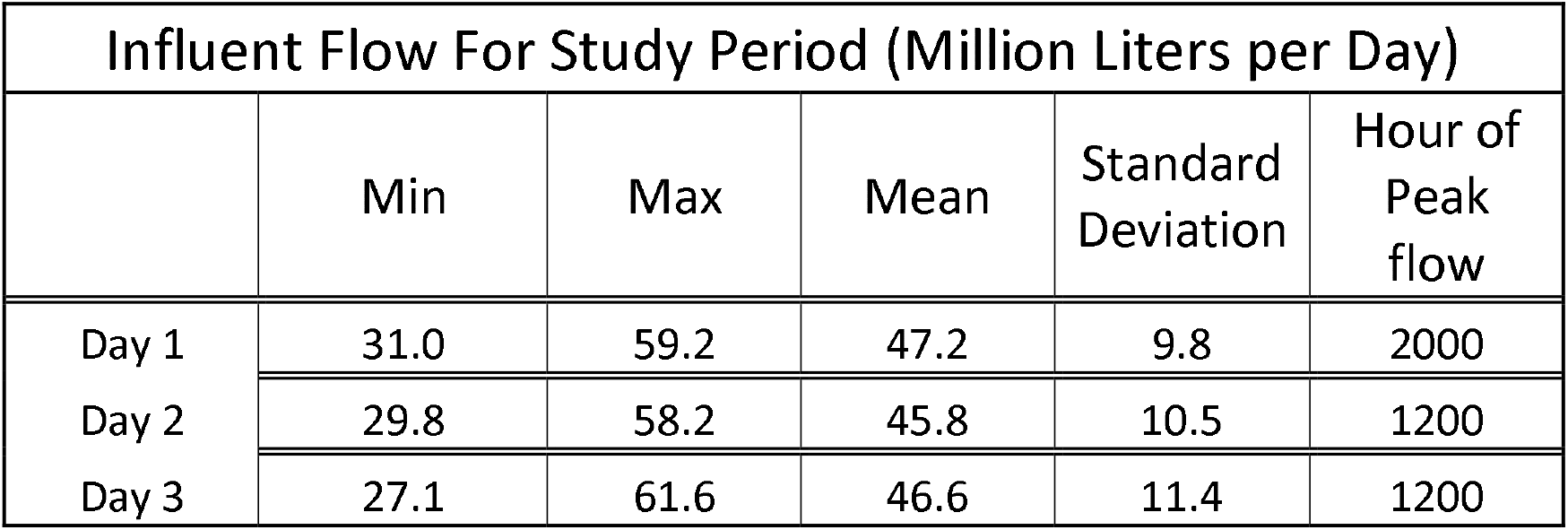
Wastewater influent flow characteristics for each of the three days during which samples were collected.

| Influent Flow For Study Period (Million Liters per Day) |  |  |  |  |  |
| --- | --- | --- | --- | --- | --- |
|  | Min | Max | Mean | Standard<br>Deviation | Hour of<br>Peak<br>flow |
| Day 1 | 31.0 | 59.2 | 47.2 | 9.8 | 2000 |
| Day 2 | 29.8 | 58.2 | 45.8 | 10.5 | 1200 |
| Day 3 | 27.1 | 61.6 | 46.6 | 11.4 | 1200 |

### 3.2 SARS-CoV-2 Concentration and Variability

All three assays used for this study (N1, N2, N3) yielded positive results for every raw wastewater influent sample. All three final effluent samples were below the limit of detection (LOD = 24, 40, and 30 copies 100 mL for N1, N2, and N3 assays, respectively). For composite samples, concentrations of all assays ranged from 580 – 1380 copies 100 Ml^-1^, with a mean of 900 and standard deviation of 215 copies 100 mL^-1^, showing good agreement across the three days (Figure 1). Similarly, composite samples showed relatively low variability within (largest range = 490 copies 100 mL^-1^) and between assays (largest range = 580 copies 100 mL^-1^) for a given day (Table 2). Grab sample concentration variability was also low, ranging from 25 to 1100 copies 100 mL for all samples collected (Table 3) with a coefficient of variation (CV) of 68.5%. Grab sample concentrations showed good agreement across assays as means, minima, and maxima were each in the same respective order of magnitude (Table 3). Examining the association between each pair of assays showed a positive monotonic relationship for all combinations, with Spearman coefficients ranging 0.72-0.90 (Figure 2). Comparing results by day for all assays showed similarly low variability with the greatest difference in any two daily mean concentrations of 114.8 copies 100 mL^-1^ (Table 3).

**Table 2.** SARS-CoV-2 RNA concentrations for each 24-hour, flow-weighted composite sample by assay.

| SARS-CoV-2 Composite Sample Concentration<br>(copies 100mL<sup>-1</sup>) |  |  |  |  |
| --- | --- | --- | --- | --- |
| Date | Composite | N1 | N2 | N3 |
| 5/2/2020 | 1 | 860 | 890 | 890 |
| 5/3/2020 | 2 | 800 | 1380 | 1010 |
| 5/4/2020 | 3 | 580 | 910 | 780 |

**Table 3.** SARS-CoV-2 grab sample RNA concentration descriptive statistics by assay and sample day.

| SARS-CoV-2 Grab Sample<br>Concentration (copies 100 mL<sup>-1</sup>) |  |  |  |  |
| --- | --- | --- | --- | --- |
| By Assay |  |  |  |  |
|  | N1 | N2 | N3 | Overall |
| Min | 50 | 90 | 140 | 50 |
| Max | 2200 | 2000 | 2100 | 1100 |
| Mean | 608 | 848 | 768 | 741 |
| St. Dev | 501 | 500 | 506 | 508 |
| By Day |  |  |  |  |
|  | Day 1 | Day 2 | Day 3 | Overall |
| Min | 220 | 50 | 110 | 50 |
| Max | 2200 | 2100 | 1800 | 1100 |
| Mean | 759 | 790 | 675 | 726 |
| St. Dev | 456 | 571 | 497 | 502 |

**Figure 1.**
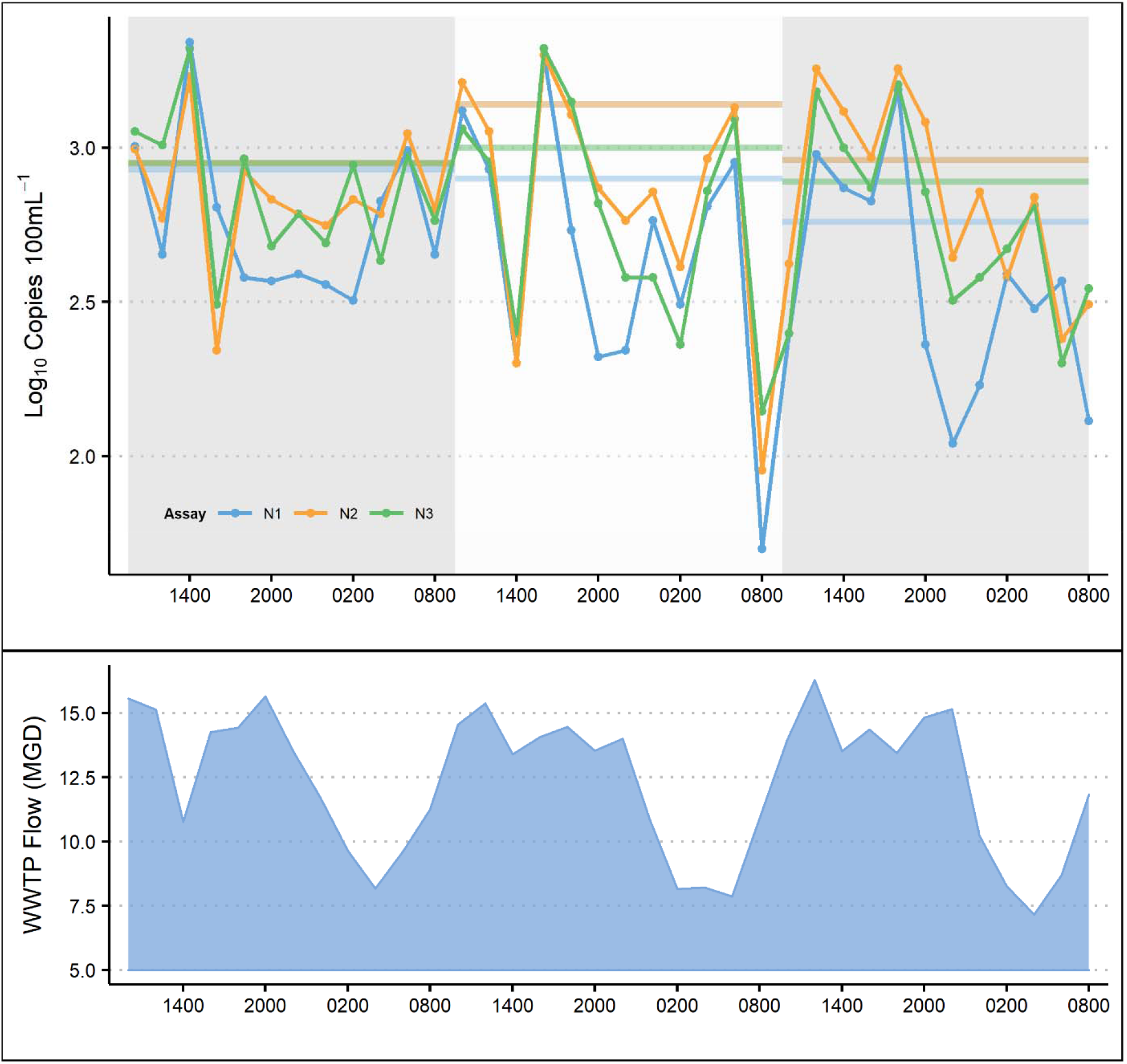
Wastewater log10 SARS-CoV-2 concentrations (copies 100 mL-1). Grab sample concentrations are denoted by dots with each color representing an assay (N1, N2, N3). Grey and white shaded areas denote the timeframe for three discrete 24hr flow-weighted composites. Horizontal lines denote concentrations for each composite sample. Influent flow is plotted in the lower panel.

**Figure 2.**
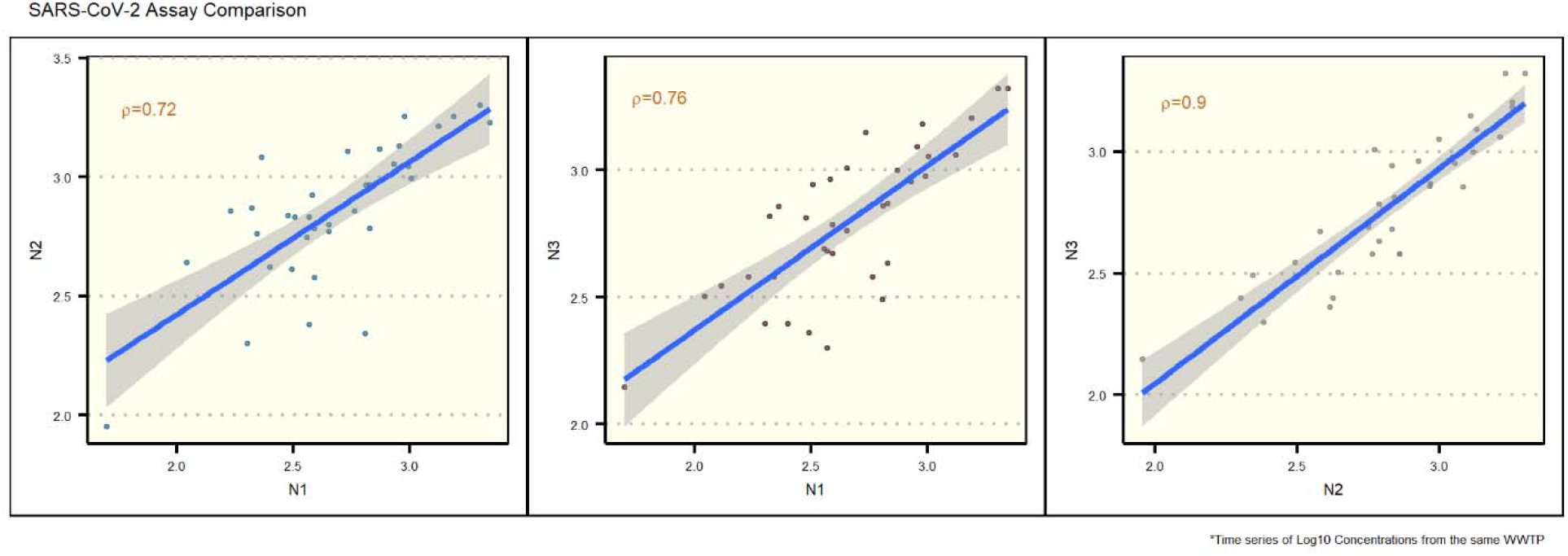
Associations between SARS-CoV-2 assays (N1, N2, N3). X and Y axes show log_10_ concentrations for each assay. Lines represent linear associations between assays, shaded areas denote standard error for each regression line. Spearman correlation coefficients are listed in orange on each plot.

Grab sample concentrations showed good agreement with corresponding composite concentrations (Figure 1), with a mean deviation of 159 copies 100 mL^-1^ between a grab sample and its associated composite. Over half (55%) of the total number of grab samples (59/108) had concentrations which were within 50% of their respective composite. The discrepancy between grab and composite concentrations, regardless of magnitude, showed grabs at lower concentration than the corresponding composite for 69% of samples (75/108) (Figure 1). These drops in virus concentration were not concurrent with times of lowest influent flow but seemed to lag by approximately 4-6hrs (Figure 1). This pattern may be influenced by the number and density of COVID-19 infections in the region. In a case with few infected individuals the viral signal would be sporadic in the daily flow. Conversely, if a catchment area were highly impacted by infections, the virus signal in wastewater would be less variable and minimally influenced by changes in flow. The ABTP catchment could have a high enough infection density to consistently detect a wastewater signal, but not so ubiquitous that the signal is entirely unimpacted by diurnal cycles in flow. Additionally, the grab samples with the extreme low concentrations were affected most by inhibition. For example, the sample with the lowest concentration (N1 = 50 copies 100mL^-1^) had a Hep G recovery (0.19%) approximately 2 orders of magnitude less than the meansample recovery (17%). It is possible that times of low flow have more concentrated matrix inhibitors that affect PCR more severely, thus underrepresenting concentrations. Considering this, grab samples should be collected at times that avoid early morning flow minima (e.g. 0800-1100) and the subsequent 4-6hrs dips in viral concentration, in order to reduce the likelihood of underestimating viral load to the treatment facility.

### 3.3 Viral Load and Carrier Prevalence

Wastewater N2 SARS-CoV-2 concentrations were used to calculate viral load for grab and composite samples (Figure 3). Viral loads calculated using composite sample values showed low variability between days, ranging from 4.2*10^11^ – 6.3*10^11^ copies day^-1^. Variability in daily load derived using grab sample concentrations was greater, ranging from 4.2*10^11^ – 9.3*10^11^ copies day^-1^, with a mean of 4.0*10^11^ copies day^-1^ and standard deviation of 2.3*10^11^ copies day^-1^. While the variability in grab sample concentration (CV=68.5%) and viral load calculated from grab sample concentration (CV=58.4%) are expectedly similar, the magnitude of any given deviation in viral load is increased due to the way load is derived (Equation 2). For example, the greatest difference in concentration between a grab and composite sample, within a common assay, was 1340 copies 100 mL^-1^. When viral load is calculated using this same grab and composite the difference between the two types of sample is 4.1*10^11^ copies day^-1^. For all load calculations using grab sample values, the mean deviation from the corresponding composite value was 9.3 *10^11^ copies day^-1^, or 16.6%. Data presented here demonstrate the large disparity in viral load values calculated using SARS-CoV-2 concentrations given a difference in only 2hrs between grab sample collection times. For this study grab samples more often had lower concentrations than the corresponding composite, thus there is a higher likelihood of underestimating concentrations when collecting grabs. Viral concentration data which are biased low will affect downstream calculations made using these data, such as estimates of viral load and carrier prevalence in the catchment population. If these metrics are used to inform a public health response it is critical that they do not systematically underestimate the extent of COVID-19 infections in the community.

**Figure 3.**
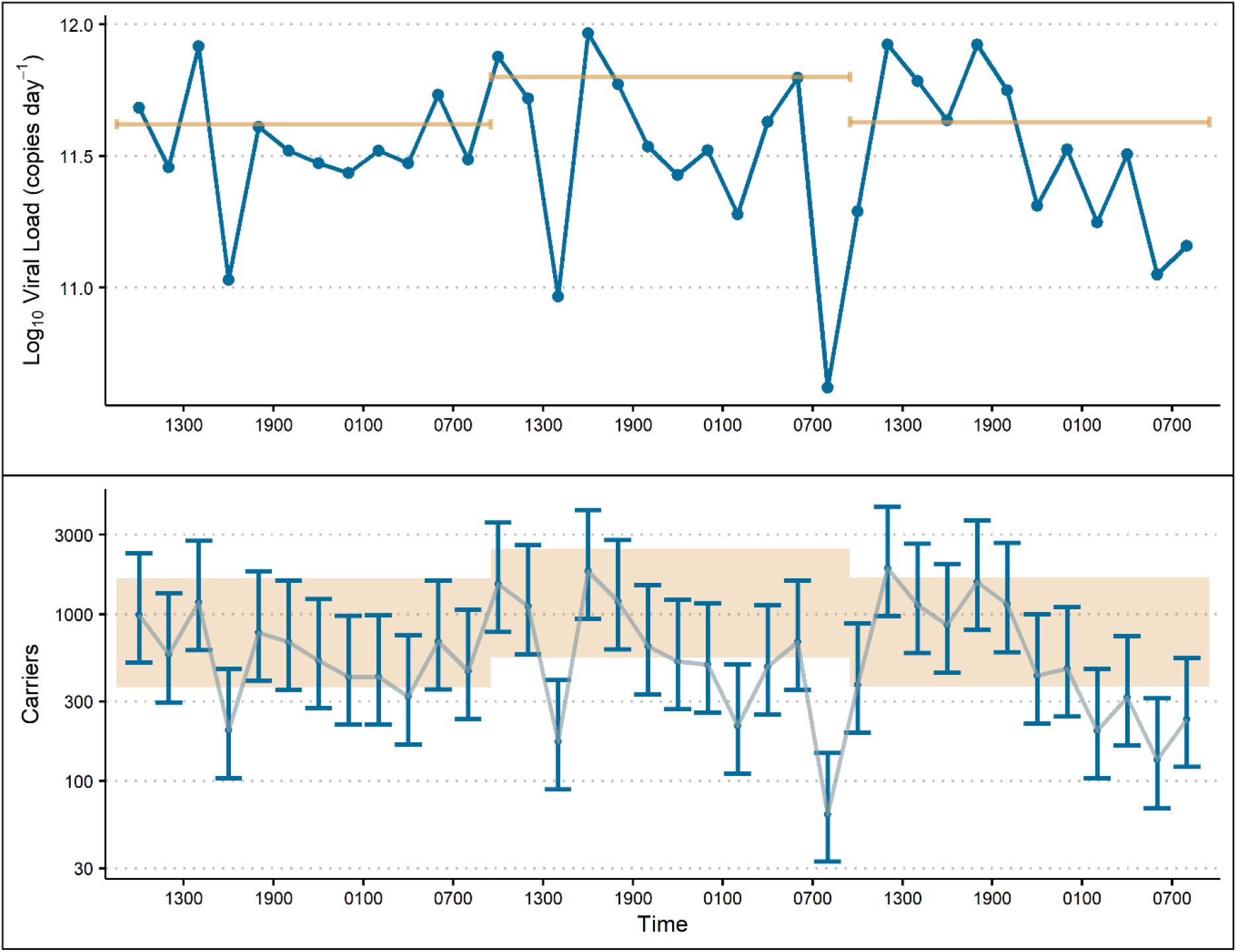
Wastewater SARS-CoV-2 load and carrier prevalence estimates for the 72-hour study period. For the upper panel, load (log_10_ copies) calculated using grab sample concentrations are denoted by blue dots while load from 24-hour composite concentrations are denoted by horizontal orange lines. In the lower panel, prevalence of SARS-CoV-2 infected carriers is estimated using Monte Carlo simulation. Estimates derived using grab sample concentrations are denote by blue dots (median number of carriers per simulation) with error bars indicating the 10^th^ and 90^th^ percentile range in estimates. Shaded areas indicate the 10^th^ and 90^th^ percentile range of carrier estimates calculated using 24-hour composite samples.

SAR-CoV-2 concentrations in wastewater can also be used to estimate the prevalence of carriers in a catchment population (Equation 3). Currently there is considerable uncertainty around the viral shedding rate in feces of people infected with COVID-19. A widely reference paper by Wölfel et al. (2020) examining nine clinical cases found concentrations of SARS-CoV-2 viral RNA in stool ranging from below the limit of detection (100 copies g^-1^) to 7.1*10^8^ copies g^-1^. Furthermore, sensitivity analysis of a previous carrier prevalence model highlights the high susceptibility to the shedding rate variability, increasing the error associated with resulting estimates. However, as viral shedding rate is more fully described, carrier estimates could become increasingly important, given the potential public health value in generating a reliable estimate of changes in infection rates in a catchment. Results of this study highlight the importance of collecting a sample that is representative of SARS-CoV-2 concentrations in wastewater, as subsequent viral load and carrier estimates are based on this value. As with viral concentration and viral load data, variability was low in carrier estimates for composite samples with median values of 714, 1074, 720 infected individuals (Figure 3). When including the 10 and 90 percentile results, estimates ranged from 369 to 2511 carriers in the catchment as estimated from composite samples. Carrier estimates based on grab samples were more variable, with 10^th^ to 90^th^ percentile model estimates ranging from 33 to 4408 carriers, and median estimates ranging from 63-1855 carriers. The median carrier estimate from 23 of 36 grab samples fell within the 10^th^-90^th^ percentile range for the corresponding composite. Of the 13 grabs for which the median carrier estimate was outside of the composite estimate 10^th^ – 90^th^ range, 12 were below the composite estimate range. Because these calculations are based on viral concentration it was expected that estimates from grabs would more often be lower than estimates made using composite concentrations. For these data, the potential underestimation of median carrier prevalence due to collecting a grab sample rather than a composite could be as large as 1011 people, based on the minimum median carrier estimate (63) and corresponding composite estimate (1074) (Figure 3). That discrepancy in estimated carriers has practical implications if WBE is used as a component of the public health response to the SARS-CoV-2 pandemic. Choosing an appropriate sampling scheme can minimize potential bias introduced into these estimates by accurately characterizing viral concentration. If replication in other studies shows that grab samples reliably underestimate viral concentration, then either composite sampling or grab samples targeting the expected peak viral concentration should be employed to reduce the likelihood of generating data which are biased low.

### 3.4 Limitations and Future Work

One important consideration for using WBE to examine viral trends during a pandemic is the heterogenous and dynamic nature of the spread of infections. Epidemiological work has shown that, particularly during the early stages of pathogen spread, rates of infection are not uniform but rather clustered in localized hotspots often driven by importation of cases (Bajardi et al., 2009) and the disproportionate effects of “superspreading” events (Lipsitch, et al., 2003). Interpreting WBE data is also confounded by transient use of the sewerage system from people who may be infected but do not live in the catchment area, e.g. tourists or people who commute to a different area for work. Restrictions such as stay-at-home orders and the subsequent reopening of cities add further complexity to the characteristics of viral spread in a community. As a result, extrapolation of findings from one catchment to the surrounding region are not often appropriate. Therefore, data and patterns presented here pertain to this specific catchment over a 3-day period, and do not easily extend to other areas or timeframes. To address this, we suggest a surveillance approach to WBE, monitoring multiple catchments on a routine basis to characterize trends specific to a region over time. As noted, variability in influent concentration change as density of cases increase or decrease within the catchment. Calculations using influent flow, such as viral load and carrier prevalence, will also be influenced by daily and seasonal changes in influent flow volume as well as short term increases due to wet weather. Regular monitoring of facilities reduces some uncertainty by establishing a context for changes in viral loading. Though estimating carrier prevalence remains challenging due to uncertainty around viral shedding rates, tracking viral load from a catchment over time may be sufficient to gain insight into community-level trends.

## 4. Conclusions

- SARS-CoV-2 concentration variability in wastewater was low (CV=68.5%) for grab samples collected every 2 hours for 72 hours
- The three CDC recommended assays (N1, N2, N3) were highly correlated, with the strongest association observed between N2 and N3 (Spearman’s rho = 0.9)
- Grab samples showed good agreement with corresponding 24hr flow-weighted composites collected from the same period (mean deviation = 159 copies 100 mL^-1^ between a grab sample and its associated composite)
- Discrepancies between grab samples and associated composite are scaled-up when data are used to estimate viral load and carrier prevalence
- Grab sample concentrations were lower than their respective composite for 69% (75/108) of samples, suggesting the possibility of introducing a bias towards lower concentration when using grabs

## Data Availability

Data are currently unavailable for distribution.

